# Post-marketing active surveillance of myocarditis and pericarditis following vaccination with COVID-19 mRNA vaccines in persons aged 12-39 years in Italy: a multi-database, self-controlled case series study

**DOI:** 10.1101/2022.02.07.22270020

**Authors:** Marco Massari, Stefania Spila-Alegiani, Cristina Morciano, Matteo Spuri, Pasquale Marchione, Patrizia Felicetti, Valeria Belleudi, Francesca Romana Poggi, Marco Lazzeretti, Michele Ercolanoni, Elena Clagnan, Emanuela Bovo, Gianluca Trifirò, Ugo Moretti, Giuseppe Monaco, Olivia Leone, Roberto Da Cas, Fiorella Petronzelli, Loriana Tartaglia, Nadia Mores, Giovanna Zanoni, Paola Rossi, Sarah Samez, Cristina Zappetti, Anna Rosa Marra, Francesca Menniti-Ippolito, the TheShinISS-vax|COVID Surveillance Group

**Affiliations:** National Centre for Drug Research and Evaluation, Istituto Superiore di Sanità (National Institute of Health), Rome, Italy; Department of Infectious Diseases, Istituto Superiore di Sanità (National Institute of Health), Rome, Italy; Department of post-marketing surveillance, Agenzia Italiana del Farmaco (Italian Medicines Agency), Rome, Italy; Department of Epidemiology ASL Roma 1, Lazio Regional Health Service, Rome, Italy; Business Intelligence, Data Science e Data Analysis, ARIA S.p.A., Milan, Italy; ARCS – Azienda Regionale di Coordinamento per la Salute, Udine, Italy; Veneto Tumour Registry, Azienda Zero, Padova, Italy; Department of Diagnostics and Public Health, University of Verona, Verona, Italy; Department of Health of Lombardy Region, Epidemiology Observatory, Milan, Italy; Institute of Pharmacology, Pharmacovigilance, Policlinico Universitario A. Gemelli, Catholic University of Sacred Heart, Rome, Italy; Immunology Unit, University Hospital, Verona, Italy; Direzione centrale salute, politiche sociali e disabilità, Friuli Venezia Giulia Region, Trieste, Italy; Centro Regionale di Farmacovigilanza, Friuli Venezia Giulia Region, Trieste, Italy

## Abstract

**Objectives:** To investigate the association between SARS-CoV-2 mRNA vaccines, BNT162b2 and mRNA-1273, and myocarditis/pericarditis.

**Design:** Self-Controlled Case Series study (SCCS) using national data on COVID-19 vaccination and emergency care/hospital admissions.

**Setting:** Italian Regions (Lombardia, Friuli Venezia Giulia, Veneto, Lazio).

**Participants:** 2,861,809 individuals, aged 12-39 years, vaccinated with the first doses of mRNA vaccines (2,405,759 BNT162b2 and 456,050 mRNA-1273) between 27 December 2020 and 30 September 2021.

**Main outcome measures:** First diagnosis of myocarditis/pericarditis within the study period. The incidence of events in the exposure risk periods (0-21 days from the vaccination day, subdivided in three equal intervals) for first and second dose was compared with baseline period. The SCCS model was fitted using conditional Poisson regression to estimate Relative Incidences (RI) and Excess of Cases (EC) per 100,000 vaccinated by dose, age, gender and brand.

**Results:** During the study period, 441 participants aged 12-39 years developed myocarditis/pericarditis (346 BNT162b2 and 95 mRNA-1273). During the 21-day risk interval there were 114 cases of myocarditis/pericarditis (74 BNT162b2 and 40 mRNA-1273) corresponding to a RI of 1.27 (0.87-1.85) and 2.16 (1.50-3.10) after first and second dose, respectively.

An increased risk of myocarditis/pericarditis at [0-7) days was observed after first [RI=6.55; 95% Confidence Interval (2.73-15.72); EC per 100,000 vaccinated=2.0 (1.5-2.3)] and second dose [RI=7.59 (3.26-17.65); EC=5.5 (4.4-5.9)] of mRNA-1273 and after second dose of BNT162b2 [RI=3.39 (2.02-5.68); EC=0.8 (0.6-1.0)]. In males, an increased risk at [0-7) days was observed after first [RI=12.28, 4.09-36.83; EC=3.8 (3.1-4.0)] and second dose [RI=11.91 (3.88-36.53); EC=8.8 (7.2-9.4)] of mRNA-1273 and after second dose of BNT162b2 [RI=3.45 (1.78-6.68); EC=1.0 (0.6-1.2)]. In females, an increased risk at [0-7) days was observed after second dose of BNT162b2 [RI=3.38 (1.47-7.74); EC=0.7 (0.3-0.9)]. At [0-7) days an increased risk following second dose of BNT162b2 was observed in the 12-17 years old [RI=5.74, (1.52-21.72); EC=1.7 (0.7-1.9)] and in 18-29 years old [RI=4.02 (1.81-8.91); EC=1.1 (0.6-1.3)]. At [0-7) days an increased risk after first [RI=7.58 (2.62-21.94); EC=3.5 (2.4-3.8)] and second [RI=9.58 (3.32-27.58); EC=8.3 (6.7-9.2)] dose of mRNA-1273 was found in 18-29 years old and after first dose in 30-39 years old [RI=6.57 (1.32-32.63); EC=1.0 (0.3-1.1)].

**Conclusions:** This population-based study indicates that mRNA vaccines were associated with myocarditis/pericarditis in the population younger than 40 years, whereas no association was observed in older subjects. The risk increased after the second dose and in the youngest for both vaccines, remained moderate following vaccination with BNT162b2, while was higher in males following vaccination with mRNA-1273. The public health implication of these findings should be weighed in the light of the overall efficacy and safety profile of both vaccines.

## Introduction

Intensive post-marketing surveillance of SARS-CoV-2 vaccines is ongoing worldwide to provide updated information on their effectiveness and safety, thereby supporting regulatory benefit/risk assessment. Since early phase of the global vaccination campaign, case series^1-3^ and pharmacovigilance reports^4,5^ on myocarditis and pericarditis following the COVID-19 mRNA vaccine administration were published. Both events were included as related to COVID-19 disease in the early and updated Priority List of COVID-19 Adverse events of special interest, developed by Brighton Collaboration Group and Safety Platform for Emergency vACcines (SPEAC), in order to harmonize safety assessment of COVID-19 vaccines in pre- and post-marketing setting.^6^ Moreover, as per core requirements for Risk Management Plan (RMP), they have been periodically monitored through routine pharmacovigilance activities in the Monthly Summary Safety Reports of all COVID-19 vaccines.^7^

On July 2021, the COVID-19 subcommittee of WHO Global Advisory Committee on Vaccine Safety reported that very rare cases of myocarditis and pericarditis had occurred more often in adolescents or young adults and after the second dose, especially within a few days after COVID-19 mRNA vaccines, and encouraged countries to strengthen the monitoring of myocarditis/pericarditis.^8^ At the same time, EMA’s Pharmacovigilance Risk Assessment Committee (PRAC) began an assessment on signals of myocarditis and pericarditis with Pfizer mRNA vaccine (BNT162b2) and Moderna mRNA vaccine (mRNA-1273) and concluded that both cardiac conditions can occur in very rare cases following vaccination with the COVID-19 mRNA vaccines. Thus, the Committee recommended to update the product information and the RMP for these vaccines, together with a direct healthcare professional communication to raise awareness among healthcare professionals.^9^

In October 2021, further data were available from an unpublished Nordic population-based register study on myocarditis and pericarditis in northern Europe, that prompted some public health organisations in the Nordic countries (e.g. Sweden, Finland, Norway, Iceland)^10^ either to pause the use of the mRNA-1273 or to recommend the use of the BNT162b2 rather than mRNA-1273 in younger people and/or younger males. In December 2021, the PRAC re-assessed the relevant safety signal, based on the Nordic study and on a study conducted using data from the French national health system (Epi-phare),^11^ concluding that the risk for both events is overall “very rare” (up to one in 10,000 vaccinated people) and greater in younger males. A further update of product information was recommended, while the benefit/risk was confirmed as positive for the whole indication.^12^

In line with these findings, recent published data from large population-based studies from Israel, US, UK and Denmark documented that the risks of myocarditis/pericarditis following mRNA vaccines differ by age groups, gender, and vaccine brand, with an higher risk in those younger than 40 years.^13-18^

In Italy, SARS-CoV-2 vaccines have been administered since late December 2020 and have been offered to the population according to a priority scheme, considering profession, age and health condition. Vaccination in adolescent (≥12 years) started on 31 May and 28 July 2021 for BNT162b2 and mRNA-1273, respectively.

Along with the enhanced passive surveillance of the Italian PharmacoVigilance network, an active surveillance, based on Regional health care claims databases, was set up by the Italian National Institute of Health (ISS) and the Italian Medicines Agency (AIFA) to provide real-world data on SARS-CoV-2 vaccine safety. In this paper, we present results of the first population based Italian study investigating the association between mRNA based COVID-19 vaccines (BNT162b2 and mRNA-1273) and myocarditis/pericarditis in the population of vaccinated persons aged 12-39 years, during the period 27 December 2020 and 30 September 2021.

## Methods

### Data source

The active surveillance is based on a dynamic multi-regional observational cohort. A distributed analysis framework is applied using *TheShinISS*, an R-based open-source statistical tool, developed by the National Institute of Health,^19^ that locally processes data collected and updated every four months from regional health care databases according to ad hoc, study-tailored, common data model.

Data on vaccination exposure, on hospitalization for myocarditis/pericarditis and on covariates were retrieved from several routinely collected regional healthcare claims databases:

- COVID-19 vaccination registry to identify information on administered vaccines (brand, date of administration and doses for all vaccinated subjects);
- population registry to identify information an age, gender and vital status; causes of death are not recorded in this registry;
- hospital discharge and emergency care visit databases to identify myocarditis/pericarditis events (pre and post vaccination) and information on a predefined list of potential confounders in the period preceding the vaccination;
- pharmacy claims and copayment exemptions databases to obtain information on potential confounders in the period preceding the vaccination;
- vaccination registry to identify other vaccinations (e.g. flu and pneumococcal vaccines) administered in the period pre- and post-COVID-19 vaccination;
- COVID-19 surveillance system to obtain information on Sars-Cov2 infection data and related outcomes.

### Study design

We used a Self-Controlled Case Series (SCCS) design.^20-24^ The SCCS design has emerged as a key methodology for studying the safety of vaccines and medicines. This approach only requires information from individuals who have experienced the event of interest, and automatically controls for multiplicative time-invariant confounders, even when these are unmeasured or unknown. Originally designed to analyze the association between vaccination and specific events under the key assumption that events do not influence post-event exposures, this method has been adapted to event dependent exposures, for example when occurrence of an event may preclude any subsequent exposure (SCCS method for censored, perturbed or curtailed post-event exposures).^23-25^ This is the case in observational studies of vaccines when the event of interest could be a contraindication to vaccination.

By using the adapted SCCS method for event dependent exposures, we estimated the relative incidence of myocarditis/pericarditis following pre-specified windows at risk after vaccination, in a within-person comparison of different time-periods. The method allows for the control of all time-independent characteristics of subjects. The SCCS method allows also for adjustment of potential time-varying confounders such as seasonal variation in risks.

### Study period and population

We investigated the association between mRNA based COVID-19 vaccines and subsequent onset of myocarditis/pericarditis in the population aged 12-39 years in the period 27 December 2020 - 30 September 2021 (the latest date for which outcome data were available). Regional claims data were locally transformed into a study-specific Common Data Model and locally processed using *TheShinISS*.

In the end, regional pseudonymized datasets were provided to the National Institute of Health for centralized analysis, in compliance with EU General Data Protection Regulation. Over the last two years, *TheShinISS* framework has been employed in several large scale observational studies exploring the association between several exposures and COVID-19 onset/prognosis as well as other drug and vaccine-related research topics and is currently maintained by a collaborative research network.^19,26-28^ The relational scheme of the study databases as well as *TheShinISS* elaborating process is described in Supplementary Figure 1.

Four Italian Regions (Lombardia, Veneto, Friuli Venezia Giulia, and Lazio), representing 36% of the population aged 12-39 years, contributed data of all vaccinated persons in this age group, in a period ranging from 27/12/2020 to the latest date for which data on outcomes were available, which varied across Regions: Lombardia up to 30/09/2021, Veneto up to 20/06/2021, Friuli Venezia Giulia up to 31/08/2021, and Lazio up to 16/06/2021). We included in the study all the persons aged 12-39 years who received a first dose of mRNA vaccines and were admitted to emergency care or hospital with the outcomes of interest. We excluded individuals with missing age and sex, or without information on vaccine brand at the first and/or second dose. Furthermore, we excluded individuals with a history of myocarditis or pericarditis within 365 days leading up to the start of the study period. The observation period for each case ranged from 27 December 2020 to the end of follow-up, which occurs at the time of death or at the end of Region-specific study period, whichever comes first.

### Definition of outcomes

The outcome of interest was the first diagnosis of myocarditis/pericarditis identified through emergency care and/or hospital admission occurring between 27 December 2020 and 30 September 2021 using International Classification of Disease, 9^th^ Revision, Clinical Modification (ICD-9-CM codes of myocarditis: 391.2; 398.0; 422; 429.0; ICD-9 codes of pericarditis: 391.0; 393; 420; 423.1; 423.2; 423.9).

### Definition of exposures

The exposures of interest were the first or second dose of BNT162b2 and mRNA-1273 vaccines. The exposure risk period was defined as [0-21) days after first or second dose administration (vaccination date), which included day 0, the day of vaccination. This risk period was further subdivided into pre-specified time-periods: [0-7), [7-14), and [14-21) after each exposure date. The unexposed baseline period (reference period) was defined as any time of observation out of the risk periods.

### Statistical analysis

Characteristics of the cohort of vaccinated persons and myocarditis and pericarditis cases were described by age, gender and comorbidity. Temporal timing of myocarditis or pericarditis events in relation to first/second dose vaccination dates was described by week.

The SCCS model was fitted using a conditional Poisson regression model to estimate the Relative Incidences (RI) and their 95% Confidence Intervals (CI). To handle event-dependent exposures the SCCS model was properly adjusted considering a counterfactual exposure history for any exposures arising after occurrence of an event.^20,24^ Five 45-day calendar periods were considered as time-varying covariate controlling for the seasonal effect. We also estimated the Excess of Cases (EC) per 100,000 vaccinated.^29^

We carried out subgroup analyses by age group (12-17, 18-29, 30-39 years), gender and vaccine brand (BNT162b2 and mRNA-1273). To assess the robustness of the primary analysis, the following sensitivity analyses were conducted: a) event-dependent exposures handling – in addition to the SCCS adjusted for event dependent exposures, we used the SCSS standard starting the observation time at the first and second dose, respectively; b) observation/exposure time period – we restricted the analysis to the study period from 27 December 2020 to 31 May 2021^20,30^ and to the study period from 1 June 2021 to 30 September 2021; we repeated the primary analysis excluding day 0 from the [0-7) day risk interval; c) heterologous vaccination – we carried out the primary analysis excluding individuals who received two different vaccine brands at the first and second dose or censoring individuals who received a different vaccine brand at the second dose (in the primary analysis the second dose was assumed to be of the same brand as the first one); d) SARS-CoV-2 infection – we restricted the analyses to subjects without a SARS-CoV-2 positive test before the occurrence of the event (any time) and within 10 days after the event. We conducted also an ancillary analysis reproducing the primary SCCS analysis in the vaccinated persons aged over 40 years.

The analyses were performed using R version 4.1.2 (R Core Team 2021) with SCCS package^31^ and STATA version 16.1.

## Results

Our cohort included 13,728,174 persons older than 12 years, who received COVID-19 vaccines between 27 December 2020 to 30 September 2021, of these 10,769,025 (78.4%) received mRNA vaccines.

During the study period, 5,109,231 doses of mRNA vaccines were administered to 2,861,809 persons aged 12-39 years (median age 26 years, interquartile range [19-33]; 49% females); 2,405,759 (84%) persons received BNT162b2 vaccine and 456,050 (16%) received mRNA-1273 vaccine (Table 1; definition of study covariates are reported in Supplementary Tables 1). The vaccinated persons had a mean follow-up time of 259 days (range 41-270).

**Table 1.** Characteristics of mRNA vaccinated population aged 12-39 years (n. 2,861,809) from 27 December 2020 to 30 September 2021, by vaccine brand.

|  | mRNA [n.(%)] | BNT162b2 [n.(%)] | mRNA-1273 [n.(%)] |
| --- | --- | --- | --- |
| **Number of subjects** | 2,861,809 | 2,405,759 | 456,050 |
| **Males** | 1,458,703 (51.0) | 1,214,517 (50.5) | 244,186 (53.5) |
| **Females** | 1,403,106 (49.0) | 1,191,242 (49.5) | 211,864 (46.5) |
| **Charlson index (1+)** | 37,679 (1.3) | 32,348 (1.3) | 5,331 (1.2) |
| **Hospitalizations in the last 2 years (1+)** | 294,859 (10.3) | 246,487 (10.2) | 47,372 (10.4) |
| **COVID-19 diagnosis before vaccination** | 238,667 (8.3) | 198,081 (8.2) | 40,586 (8.9) |
| **Prescriptions in the last 12 months (1+)** | 1,043,132 (36.5) | 876,976 (36.5) | 166,156 (36.4) |
| **COPD** | 163,804 (5.7) | 139,260 (5.8) | 24,544 (5.4) |
| **Chronic pulmonary disease** | 10,405 (0.4) | 8,718 (0.4) | 1,687 (0.4) |
| **Chronic kidney failure** | 5,789 (0.2) | 4,941 (0.2) | 848 (0.2) |
| **Neoplasms** | 36,087 (1.3) | 31,097 (1.3) | 4,990 (1.1) |
| **Diabetes mellitus** | 45,964 (1.6) | 38,820 (1.6) | 7,144 (1.6) |
| **Hematologic disease** | 148,429 (5.2) | 123,698 (5.1) | 24,731 (5.4) |
| **Cardiovascular and cerebrovascular diseases** | 52,539 (1.8) | 44,837 (1.9) | 7,702 (1.7) |
| **Hypertension** | 48,023 (1.7) | 40,625 (1.7) | 7,398 (1.6) |
| **Hepatopathy** | 9,669 (0.3) | 7,931 (0.3) | 1,738 (0.4) |
| **HIV** | 5,736 (0.2) | 4,404 (0.2) | 1,332 (0.3) |
| **Rheumatic diseases** | 25,906 (0.9) | 22,243 (0.9) | 3,663 (0.8) |
| **Cystic fibrosis** | 1,192 (0.04) | 1,101 (0.05) | 91 (0.02) |
| **Neurological diseases** | 116,248 (4.1) | 97,748 (4.1) | 18,500 (4.1) |
| **Peptic ulcer** | 123,509 (4.3) | 103,063 (4.3) | 20,446 (4.5) |
| **Colitis** | 11,249 (0.4) | 9,588 (0.4) | 1,661 (0.4) |
| **Celiac disease** | 22,056 (0.8) | 19,079 (0.9) | 2,977 (0.7) |
| **Infection** | 521,940 (18.2) | 437,186 (18.2) | 84,754 (18.6) |
| **Corticosteroids for systemic use** | 100,096 (3.5) | 84,220 (3.5) | 15,876 (3.5) |
| **NSAIDs use** | 26,108 (0.9) | 21,621 (0.9) | 4,487 (1.0) |
| **Estroprogestinics use** | 27,783 (1.0) | 23,282 (1.0) | 4,501 (1.0) |

n.: number; yrs: years; COPD: Chronic Obstructive Pulmonary Disease; HIV: Human Immunodeficiency Virus; NSAIDs: Non Steroidal Anti-Inflammatory Drugs

During the study period, 441 had an emergency care and/or hospital admission related to myocarditis/pericarditis. Of these, 302 (68.5%) were males and 139 (31.5%) were females; there were 346 (78.5%) cases in those vaccinated with BNT162b2 and 95 (21.5%) in those vaccinated with mRNA-1273 (Table 2). Figure 1 describes the temporal trend of the occurrence of the events relative to vaccination date. We observed one death, for unknown cause, after 38 days following a pericarditis case that occurred 53 days after the second dose of BNT162b2 vaccine (unexposed period).

**Table 2.** Characteristics of cases of myocarditis/pericarditis (n. 441) among the mRNA vaccinated population aged 12-39 years from 27 December 2020 to 30 September 2021, by age group and vaccine brand.

|  | mRNA [n.(%)] |  |  |  | BNT162b2 [n.(%)] |  |  |  | mRNA-1273 [n.(%)] |  |  |  |
| --- | --- | --- | --- | --- | --- | --- | --- | --- | --- | --- | --- | --- |
|  | 12-39yrs | 12-17yrs | 18-29yrs | 30-39yrs | 12-39yrs | 12-17yrs | 18-29yrs | 30-39yrs | 12-39yrs | 12-17yrs | 18-29yrs | 30-39yrs |
| Number of subjects | **441** | 57 | 211 | 173 | **346** | 46 | 154 | 146 | **95** | 11 | 57 | 27 |
| Males | **302 (68.5)** | 43 (75.4) | 152 (72.0) | 107 (61.8) | **232 (67.1)** | 33 (71.7) | 105 (68.2) | 94 (64.4) | **70 (73.7)** | 10 (90.9) | 47 (82.5) | 13 (48.1) |
| Females | **139 (31.5)** | 14 (24.6) | 59 (28.0) | 66 (38.2) | **114 (32.9)** | 13 (28.3) | 49 (31.8) | 52 (35.6) | **25 (26.3)** | 1 (9.1) | 10 (17.5) | 14 (51.9) |
| Charlson index (1+) | **53 (12.0)** | 4 (7.0) | 19 (9.0) | 35 (20.2) | **46 (13.3)** | 2 (4.3) | 14 (9.1) | 30 (20.5) | **7 (7.4)** | 2 (18.2) | 5 (8.8) | 5 (18.5) |
| Hospitalizations in the last 2 years (1+) | **188 (42.6)** | 18 (31.6) | 84 (39.8) | 86 (49.7) | **154 (44.5)** | 13 (28.3) | 67 (43.5) | 74 (50.7) | **34 (35.8)** | 5 (45.5) | 17 (29.8) | 12 (44.4) |
| COVID-19 diagnosis before vaccination | **60 (13.6)** | 9 (15.8) | 25 (11.8) | 30 (17.3) | **48 (13.9)** | 1 (2.2) | 21 (13.6) | 26 (17.8) | **12 (12.6)** | 8 (72.7) | 4 (7.0) | 4 (14.8) |
| Prescriptions in the last 12 months (1+) | **307 (69.6)** | 34 (59.6) | 140 (66.4) | 133 (76.9) | **246 (71.1)** | 27 (58.7) | 106 (68.8) | 113 (77.4) | **61 (64.2)** | 7 (63.6) | 34 (59.6) | 20 (74.1) |
| COPD/Asthma | **32 (7.3)** | 4 (7.0) | 15 (7.1) | 13 (7.5) | **28 (8.1)** | 3 (6.5) | 14 (9.1) | 11 (7.5) | **4 (4.2)** | 1 (9.1) | 1 (1.8) | 2 (7.4) |
| Chronic pulmonary disease | **16 (3.6)** | 3 (5.3) | 2 (0.9) | 12 (6.9) | **15 (4.3)** | 2 (4.3) | 2 (1.3) | 11 (7.5) | **1 (1.1)** | 1 (9.1) | 0 | 1 (3.7) |
| Chronic kidney failure | **9 (2.0)** | 7 (12.3) | 2 (0.9) | 9 (5.2) | **7 (2.0)** | 7 (15.2) | 2 (1.3) | 7 (4.8) | **2 (2.1)** | 0 | 0 | 2 (7.4) |
| Neoplasms | **16 (3.6)** | 3 (5.3) | 5 (2.4) | 10 (5.8) | **14 (4.0)** | 1 (2.2) | 5 (3.2) | 8 (5.5) | **2 (2.1)** | 2 (18.2) | 0 | 2 (7.4) |
| Diabetes mellitus | **8 (1.8)** | 4 (7.0) | 1 (0.5) | 7 (4.0) | **4 (1.2)** | 4 (8.7) | 1 (0.6) | 4 (2.7) | **4 (4.2)** | 0 | 0 | 3 (11.1) |
| Hematologic disease | **50 (11.3)** | 5 (8.8) | 19 (9.0) | 32 (18.5) | **43 (12.4)** | 4 (8.7) | 13 (8.4) | 26 (17.8) | **7 (7.4)** | 1 (9.1) | 6 (10.5) | 6 (22.2) |
| Cardiovascular and cerebrovascular diseases | **170 (38.5)** | 18 (31.6) | 78 (37.0) | 74 (42.8) | **138 (39.9)** | 13 (28.3) | 61 (39.6) | 64 (43.8) | **32 (33.7)** | 5 (45.5) | 17 (29.8) | 10 (37.0) |
| Hypertension | **82 (18.6)** | 4 (7.0) | 27 (12.8) | 51 (29.5) | **66 (19.1)** | 1 (2.2) | 21 (13.6) | 44 (30.1) | **16 (16.8)** | 3 (27.3) | 6 (10.5) | 7 (25.9) |
| Hepatopathy | **2 (0.5)** | 1 (1.8) | 1 (0.5) | 2 (1.2) | **1 (0.3)** | 1 (2.2) | 1 (0.6) | 1 (0.7) | **1 (1.1)** | 0 | 0 | 1 (3.7) |
| HIV | **2 (0.5)** | 1 (1.8) | 1 (0.5) | 1 (0.6) | **2 (0.6)** | 1 (2.2) | 1 (0.6) | 1 (0.7) | **0** | 0 | 0 | 0 |
| Rheumatic diseases | **23 (5.2)** | 3 (5.3) | 8 (3.8) | 15 (8.7) | **18 (5.2)** | 1 (2.2) | 5 (3.2) | 12 (8.2) | **5 (5.3)** | 2 (18.2) | 3 (5.3) | 3 (11.1) |
| Cystic fibrosis | **0** | 0 0 | 0 0 | 0 | **0** | 0 | 0 | 0 | **0** | 0 | 0 | 0 |
| Neurological diseases | **26 (5.9)** | 7 (12.3) | 9 (4.3) | 12 (6.9) | **20 (5.8)** | 3 (6.5) | 7 (4.5) | 10 (6.8) | **6 (6.3)** | 4 (36.4) | 2 (3.5) | 2 (7.4) |
| Peptic ulcer | **152 (34.5)** | 13 (22.8) | 68 (32.2) | 71 (41.0) | **122 (35.3)** | 9 (19.6) | 52 (33.8) | 61 (41.8) | **30 (31.6)** | 4 (36.4) | 16 (28.1) | 10 (37.0) |
| Colitis | **5 (1.1)** | 2 (3.5) | 2 (0.9) | 1 (0.6) | **4 (1.2)** | 1 (2.2) | 2 (1.3) | 1 (0.7) | **1 (1.1)** | 1 (9.1) | 0 | 0 |
| Celiac disease | **1 (0.2)** | 1 (1.8) | 0 0 | 1 (0.6) | **1 (0.3)** | 1 (2.2) | 0 | 1 (0.7) | **0** | 0 | 0 | 0 |
| Infection | **155 (35.1)** | 17 (29.8) | 67 (31.8) | 71 (41.0) | **125 (36.1)** | 13 (28.3) | 53 (34.4) | 59 (40.4) | **30 (31.6)** | 4 (36.4) | 14 (24.6) | 12 (44.4) |
| Corticosteroids for systemic use | **57 (12.9)** | 4 (7.0) | 24 (11.4) | 29 (16.8) | **49 (14.2)** | 3 (6.5) | 21 (13.6) | 25 (17.1) | **8 (8.4)** | 1 (9.1) | 3 (5.3) | 4 (14.8) |
| NSAIDs use | **48 (10.9)** | 5 (8.8) | 20 (9.5) | 23 (13.3) | **40 (11.6)** | 4 (8.7) | 16 (10.4) | 20 (13.7) | **8 (8.4)** | 1 (9.1) | 4 (7.0) | 3 (11.1) |
| Estroprogestinics use | **2 (0.5)** | 1 (1.8) | 1 (0.5) | 2 (1.2) | **1 (0.3)** | 1 (2.2) | 1 (0.6) | 1 (0.7) | **1 (1.1)** | 0 | 0 | 1 (3.7) |

n.: number; yrs: years; COPD: Chronic Obstructive Pulmonary Disease; HIV: Human Immunodeficiency Virus; NSAIDs: Non Steroidal Anti-Inflammatory Drugs

**Figure 1.**
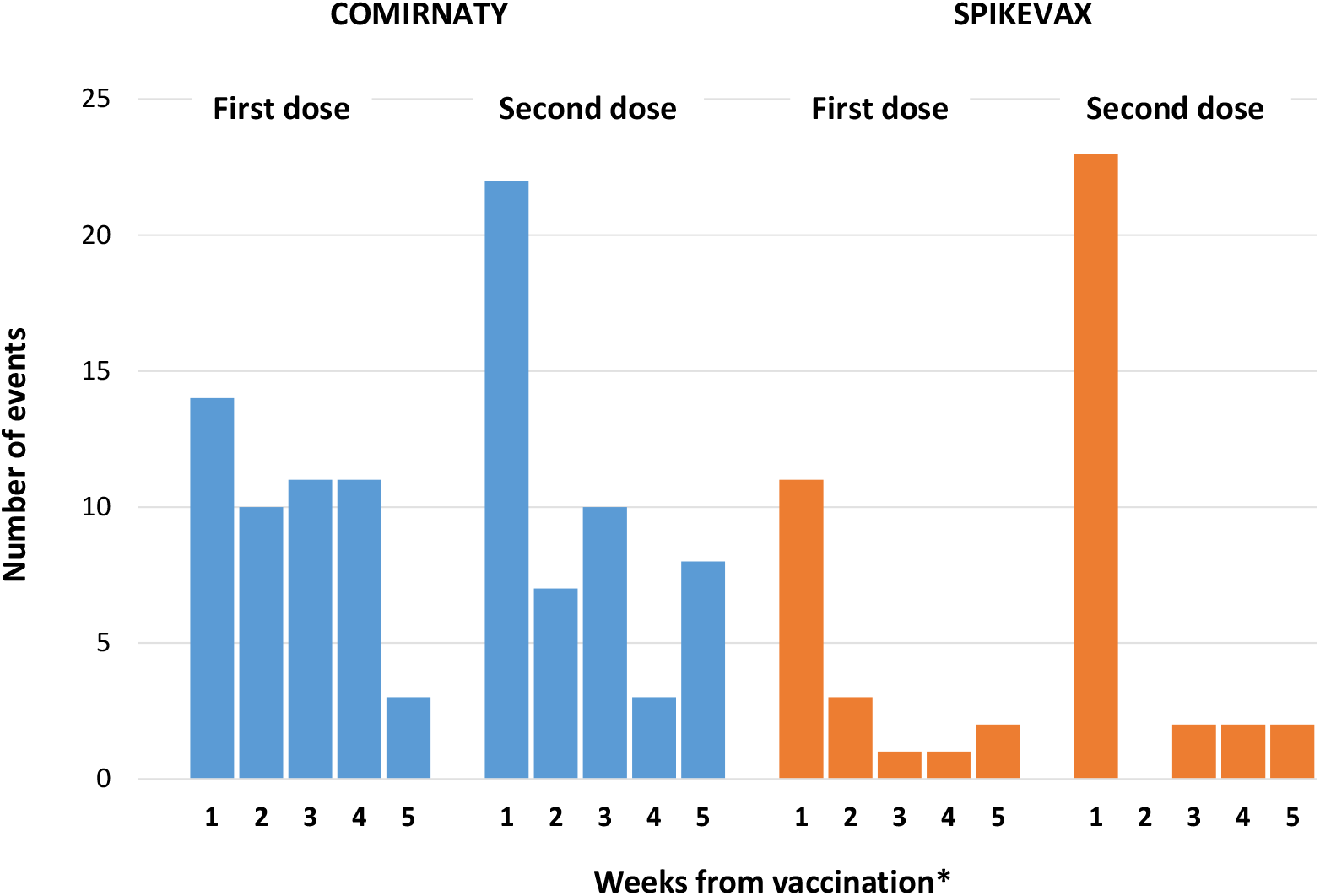
Frequency of myocarditis/pericarditis events in each week relative to the day of vaccination (the week of vaccination starts from day 0, the day of vaccination) in the vaccinated population aged 12-39 years from 27 December 2020 to 30 September 2021, by vaccine brand and dose. *only first 5 weeks from vaccination day are plotted

Tables 3 reports the results of the primary analysis from the SCCS model for the 441 cases aged 12-39 years. During the 21-day risk interval there were a total of 114 cases of myocarditis/pericarditis (74 with BNT162b2 and 40 with mRNA-1273), corresponding to a RI of 1.27 (0.87-1.85) and 2.16 (1.50-3.10) after first and second dose, respectively. The majority of these cases occurred within the [0-7) day risk period after the first or second dose administration (n. 70, 61.4%). An increased risk of myocarditis/pericarditis [0-7) days following a first dose of mRNA-1273 was observed (RI=6.55; 95% CI 2.73-15.72), while no association was found with BNT162b2. An increased risk of myocarditis/pericarditis [0-7) days was also observed following a second dose of BNT162b2 (RI=3.39; 95% CI 2.02-5.68) and mRNA-1273 (RI=7.59; 95% CI 3.26-17.65). Over the [0-7) days post-vaccination, we estimated an additional 2.0 (95% CI 1.5-2.3) myocarditis/pericarditis cases per 100,000 vaccinated persons following the first dose of mRNA-1273; following a second dose of the BNT162b2 and mRNA-1273, we estimated an additional 0.8 (95% CI 0.6-1.0) and 5.5 (95% CI 4.4-5.9) myocarditis/pericarditis cases per 100,000 vaccinated, respectively.

**Table 3.**
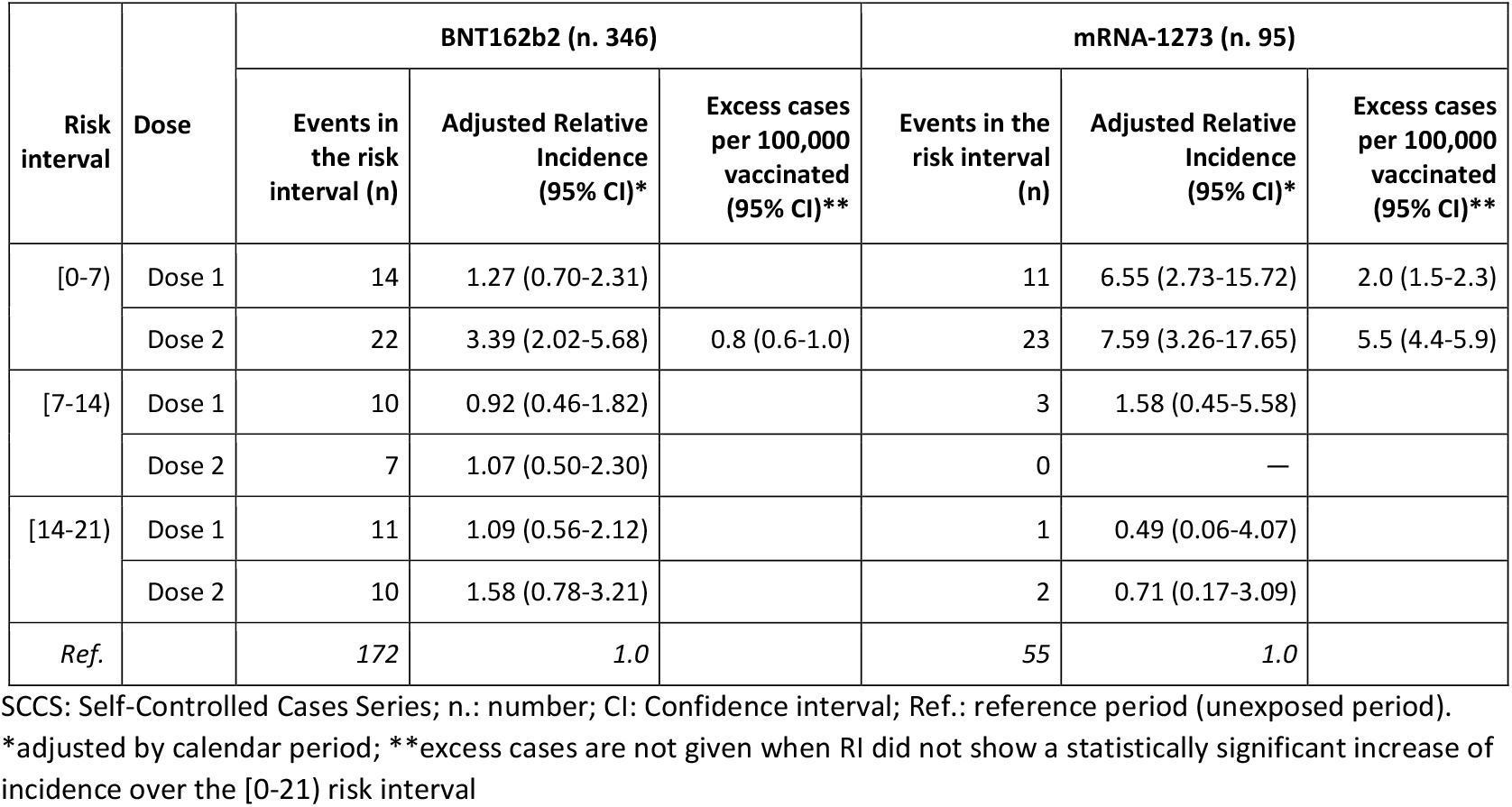
Relative incidence estimated by adjusted SCCS and excess cases per 100,000 vaccinated by vaccine brand and risk intervals: 346 myocarditis/pericarditis events in the BNT162b2 and 95 events in the mRNA-1273 vaccinated population aged 12-39 years from 27 December 2020 to 30 September 2021.

### Subgroup analysis by gender and age group

Table 4 and Figure 2 show the Relative Incidences and EC in the [0-7) day risk period by age and gender (Supplementary Tables 2-11, Supplementary Figure 2).

**Table 4.**
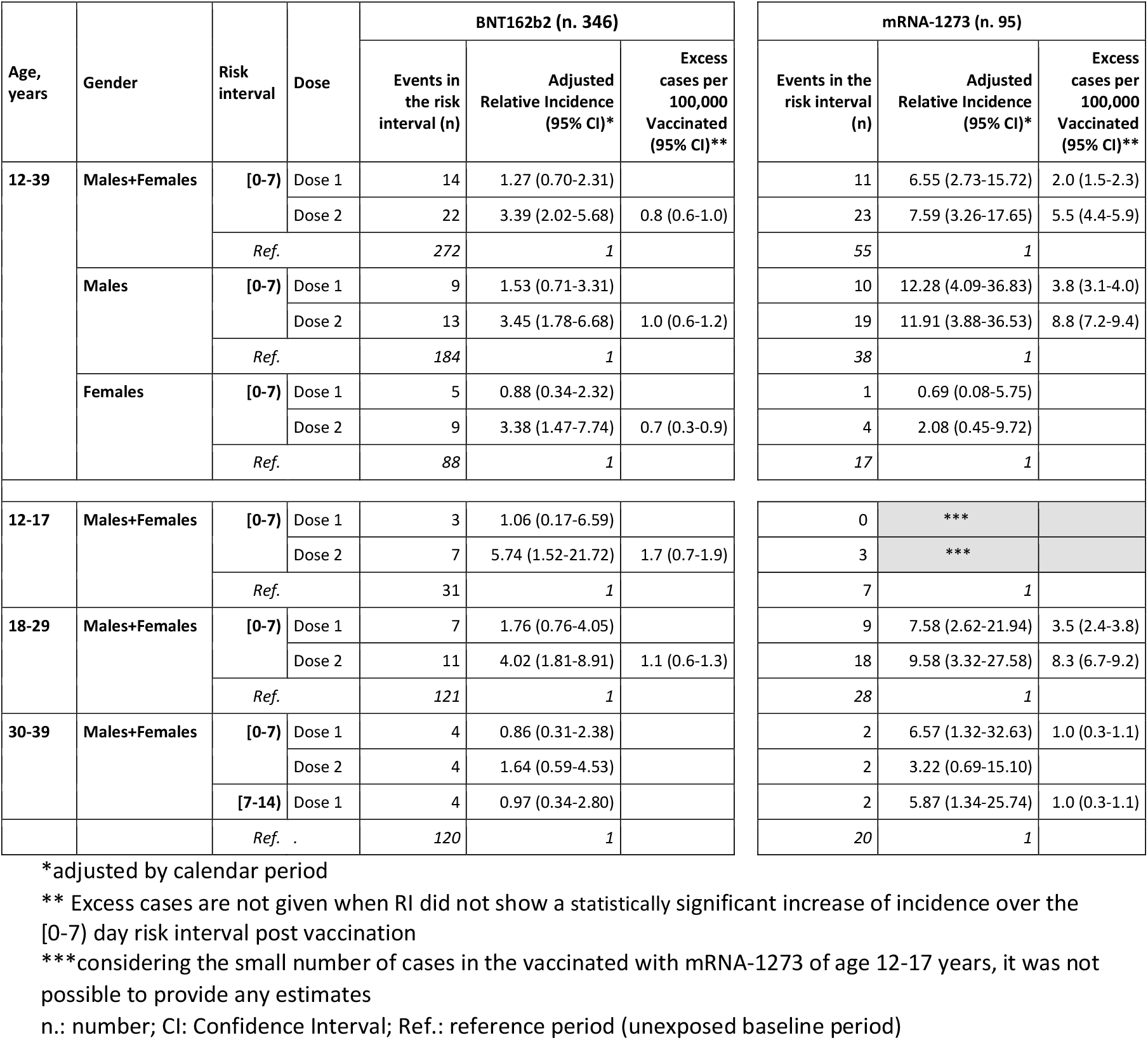
Relative Incidence and excess of cases per 100,000 vaccinated in the [0-7) risk period after mRNA vaccination in the vaccinated population aged 12-39 years from 27 December 2020 to 30 September 2021 by gender, age group and vaccine brand.

**Figure 2.**
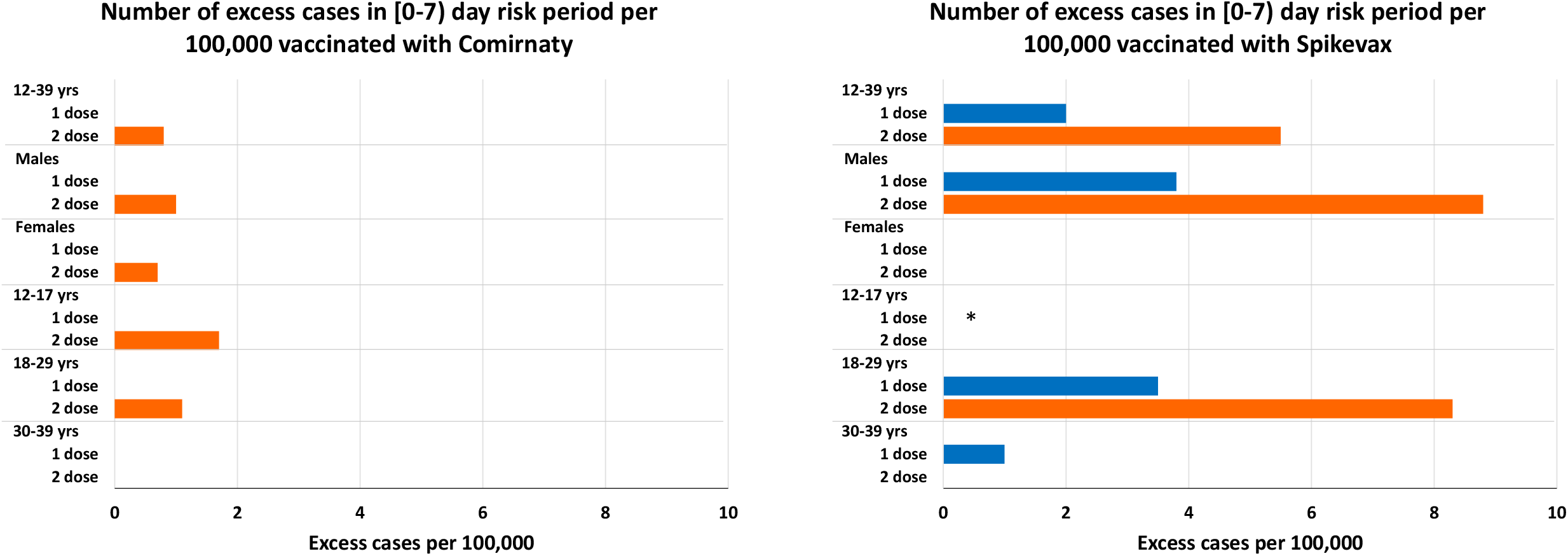
Excess of cases per 100,000 vaccinated in the [0-7) days risk period following BNT162b2 and mRNA-1273 vaccination in the vaccinated population aged 12-39 years from 27 December 2020 to 30 September 2021, by gender, age group and dose (first dose blue, second dose orange). yrs: years; *Considering the small number of cases in the vaccinated with mRNA-1273 of age 12-17 years, it was not possible to provide any estimate Excess cases are not given when RI did not show a statistically significant increase of incidence over the [0-7) day risk interval post vaccination

In males, the risk of myocarditis/pericarditis increased in the [0-7) days following a first dose of mRNA-1273 (RI=12.28; 95% CI 4.09-36.83), and following a second dose of BNT162b2 (RI=3.45; 95% CI 1.78-6.68) and mRNA-1273 (RI=11.91; 95% CI 3.88-36.53). In females, we found an increased risk of myocarditis/pericarditis [0-7) days following a second dose of BNT162b2 (RI=3.38; 95% CI 1.47-7.74), while no association was observed with mRNA-1273.

In males, we estimated an additional 3.8 (95% CI 3.1-4.0) EC per 100,000 in the [0-7) days following a first dose of mRNA-1273, and an additional 1.0 (95% CI 0.6-1.2) and 8.8 (95% CI 7.2-9.4) EC per 100,000 in the [0-7) days following a second dose of BNT162b2 and mRNA-1273, respectively. In females, we estimated an additional 0.7 (95% CI 0.3-0.9) EC per 100,000 in the [0-7) days following a second dose of BNT162b2.

Analyzing data by age group, we estimated an increased risk of myocarditis/pericarditis [0-7) days following the second dose of BNT162b2 (RI=5.74; 95% CI 1.52-21.72) in those aged 12-17 years. Number of events were insufficient to estimate associations with mRNA-1273. There were, however, three events in the [0-7) day interval compared to one event in the remaining risk intervals after the second dose. In the 18-29 year age group, we observed an increased risk of myocarditis/pericarditis [0-7) days following a first and second dose of mRNA-1273, respectively (RI=7.58; 95% CI 2.62-21.94) and (RI=9.58; 95% CI 3.32-27.58), and following the second dose of BNT162b2 (RI=4.02; 95% CI 1.81-8.91). In the age group 30-39 years, we found an increased risk of myocarditis/pericarditis [0-7) days (RI=6.57; 95% CI 1.32-32.63) and [7-14) days (RI=5.87; 95% CI 1.34-25.74) following the first doses of mRNA-1273, while no association was observed with BNT162b2.

In the age group 12-17 years, we estimated an additional 1.7 (95% CI 0.7-1.9) EC per 100,000 in the [0-7) days following a second dose of BNT162b2. In the age group 18-29 years, we estimated an additional 3.5 (95% CI 2.4-3.8) EC per 100,000 in the [0-7) days following a first dose of mRNA-1273; an additional 1.1 (95% CI 0.6-1.3) and 8.3 (95% CI 6.7-9.2) EC per 100,000 in the [0-7) days following a second dose of BNT162b2 and mRNA-1273, respectively. In the age group 30-39 years, we estimated an additional 1.0 (95% CI 0.3-1.1) EC per 100,000 both in the [0-7) days and [7-14) days following the first dose of mRNA-1273, respectively.

### Sensitivity and ancillary analyses

All sensitivity analyses were consistent with the main results of the study (Supplementary Table 12). The sensitivity analysis that was conducted to highlight the potential effect of notoriety bias (by restricting the observation period before and after 31 May 2021) indicated that RIs estimates and confidence intervals are largely overlapping, even though we cannot rule out a slight inflation of the estimates in the second period.

The ancillary analysis on 2,050 cases aged over 40 years (BNT162b2 n. 1,759; mRNA-1273 n. 291) showed no association for BNT162b2 and mRNA-1273 after seven days following the first dose [(RI=0.59 (0.42-0.82) and RI= 0.56 (0.23-1.36) and the second dose [(0.84 (0.61-1.16) and RI= 1.11 (0.57-2.17)] (Supplementary Table 13).

## Discussion

### Principal findings

This first Italian large population-based study covering about 3 million of vaccinated persons aged 12-39 years in Italy found an association between myocarditis/pericarditis within a week following each dose of mRNA vaccines.

The risk of myocarditis/pericarditis is particularly higher after seven days following the first or second dose of mRNA-1273 vaccine in the overall population. Subgroup analysis by gender suggested that the increased risk was present only in males after both the first and second dose with 3.8 and 8.8 EC per 100,000 vaccinated, respectively. Stratifying by age, greater risks were found in those aged 18-29 years with EC of 3.5 and 8.3 per 100,000 following the first and the second doses, respectively. In the age group 12-17 years, the number of events were insufficient for risk estimate.

We also observed an association between BNT162b2 and myocarditis/pericarditis, but only in the seven days following the second dose, where the risks remain similar between males and females with 1.0 and 0.7 EC per 100,000 vaccinated, respectively. In the age groups 12-17 years and 18-29 years, where the increased risks were confined, the estimated EC were 1.7 and 1.1 per 100,000 vaccinated, respectively.

Vaccine-associated acute myocarditis is generally attributable to allergic/hypersensitivity reactions as observed in other vaccines.^32^ However, the pathophysiology of myocarditis and pericarditis associated to mRNA vaccines is not clearly understood and different mechanisms have been postulated. Molecular mimicry between the spike protein and self-antigens,^33^ trigger of preexisting dysregulated immune pathways, immune response to mRNA^34^ or dysregulated cytokine expression^35^ have been proposed.

Our results on the increased risk in the seven days after each dose are consistent with the onset of viral myocarditis symptoms often reported in the first week from the infection.^32,36^

Moreover, it has been postulated that a very high antibody response to mRNA vaccines in predisposed young people may elicit an uncontrolled inflammatory response similar to multisystem inflammatory syndrome observed in children (MIS-C) with SARS-CoV-2 infection.^37^ To date, no clear evidence is available and further studies are needed to clarify which is the exact mechanism of mRNA vaccines-related myocarditis and pericarditis.

Furthermore, our observation on the increased risk in young males resembles classical epidemiological features of myocarditis due to other causes,^38^ included COVID-19 related myocarditis,^39^ but the exact role of age and sex is still unclear. In a recent review, a possible effect of sex hormones in immune response is summarized, with a possible role of testosterone by a combined mechanism of inhibition of anti-inflammatory cells and commitment to a Th1-type immune response in male and of inhibitory effects of estrogen on proinflammatory T cells in female.^40^

### Comparison with related studies

In line with a previous US study^15^ we identified an association between mRNA vaccines and myocarditis/pericarditis in individuals younger than 40 years within the 0-7 day period following the first and the second doses.

Our results are also consistent with observational studies which documented markedly increased risk of myocarditis in England,^16^ and myocarditis or myopericarditis in Denmark^18^ in the population vaccinated with mRNA-1273. Specifically, in the Danish study it was reported a strong association between mRNA-1273 and myocarditis or myopericarditis within 28 days from vaccination (Hazard Ratio, HR=5.24; 95% CI 2.47-11.12) with an estimated of 57 EC per 1,000,000 vaccinated. The UK study also suggested a strong association within the 1-28 days after first and second mRNA-1273 dose (IRR=3.89; 95% CI 1.60-9.44; and 20.71, 95% CI 4.02-106.68; respectively) corresponding to 8 and 15 EC per 1,000,000 vaccinated.^16^ A recent updated self-controlled case series analysis of English data, stratified by age and gender, also reported an higher risk in male aged less than 40 years (first dose Incidence Rate Ratio, IRR=2.34; 95% CI 1.03-5.34; second dose IRR=16.52; 95% CI 9.10-30.00) corresponding to EC of 12 and 101 per 1,000,000 respectively; a markedly increased risk was also observed in females after the second dose of mRNA-1273 (IRR=7.55; 95% CI 1.67-34.12) with 8 EC of per 1,000,000 vaccinated.^17^

Results on the association between BNT162b2 and myocarditis/pericarditis are less conclusive. We found an association in the seven days after the second dose both in males and females. Findings from Israel^14^ and England^17^ confirmed an association in adolescent and adult males younger than 40 years, but not in female participants. Particularly, the English study, including data on the third dose of BNT162b2, highlighted that in males 12-39 years the risk sequentially increased following each dose of vaccine (IRR=1.66, 3.41 and 7.60, respectively) with an EC of 3, 12 and 13 per 1,000,000 vaccinated, respectively. No association was found in females and in males older than 40 years.^17^

Conversely, a population-based study conducted in Denmark^18^, with a more stringent case definition, did not support the association between BNT162b2 and myocarditis or myopericarditis in the 28 days after vaccination, both overall and in the 12-39 year group, but an association only in females (HR=3.73; 95% CI 1.82-7.65) was found.

### Strengths and limitations

Our study strengths include the large sample size, the broad geographical distribution of the cohort and the availability of data on SARS-CoV-2 diagnosis and outcomes, comorbidities and patients’ demographic characteristics from health care databases. The large sample size (about 3 million vaccinated people aged 12-39 years) allowed to look at fine risk intervals following vaccinations and carrying several subgroup analyses. Since data were collected from routinely collected data in the claims databases, irrespective of research question, there is no potential for recall or selection bias. Another methodological strength of our study is the choice of the SCCS method modified to handle event-dependent exposures. This method properly controls for the estimate inflation occurring when vaccination is markedly delayed or cancelled (i.e. myocarditis/pericarditis is a contraindication of vaccination). Conversely, the standard SCCS method adapted by including a pre-vaccination risk period is a strategy working for short term delays.^21,23,24^

However, our study also has limitations. First, there is the possibility of notoriety bias due to overdiagnosis of cases of myocarditis/pericarditis because of the increased public and medical awareness of these potential adverse events following mRNA vaccination. Such bias is probably minimal and the effect observed in the sensitivity analysis could be partly explained by a different age profile and characteristics of the two vaccinated population before and after 31 May 2021. Second, diagnoses of myocarditis and pericarditis were retrieved from hospital discharge and emergency care visit databases, and they were not validated through the review of clinical records. For this reason, a misclassification of the outcomes that biased the association cannot be excluded. However, an over or under-representation of the ascertainment of the outcomes is unlikely, since we relied on the settings from which codes were derived i.e. emergency care and hospital care where the available facilities made it possible an accurate diagnosis, that it is known cannot be based only on clinical assessment. Third, we did not collect further information to assess the severity of the outcomes. To date, data were collected without differentiating between emergency care admission and hospital admission and length of the hospitalization was not available. Lastly, although the large sample size including about three million of vaccinated persons, due to the small number of events, it was not possible to provide robust model estimates in some subgroup analyses (i.e. mRNA-1273 in the subgroup of adolescents aged 12-17 years).

## Conclusions

This population-based study indicates that mRNA vaccines were associated with myocarditis/pericarditis in the population younger than 40 years, whereas no association was observed in older subjects. According to our results the risk increased after the second dose and in the youngest for both vaccines, remained moderate following vaccination with BNT162b2, while was higher in males following vaccination with mRNA-1273. However, vaccine-associated risks should always be evaluated in the light of their efficacy in preventing serious COVID-19 disease and death.

Further monitoring of data from this active surveillance is needed to evaluate the relationship between mRNA vaccines and myocarditis/pericarditis by age within gender, including population of children (5-11 years old) and the effect of third dose (booster dose).

## Data Availability

Due to regional and national data privacy regulations, individual-level data cannot be shared openly. Aggregated or jittered data will be shared on reasonable request to the corresponding author (CM).

## Ethics statements

### Ethics approval

This study was approved by the National Unique Ethics Committee for the evaluation of clinical trials of medicines for human use and medical devices for patients with COVID-19 of the National Institute for Infectious Diseases “Lazzaro Spallanzani” in Rome (n. 335, 17/05/2021 and n. 399, 02/09/2021).

## Acknowledgements

We would like to thank Gianpaolo Scalia Tomba, Giuseppe Traversa, Maria Paola Trotta, Nicola Magrini and Patrizia Popoli for their useful suggestions and *TheShinISS-vax COVID Surveillance Group:* Francesca Menniti Ippolito, Roberto Da Cas, Ilaria Ippoliti, Marco Massari, Cristina Morciano, Paola Ruggeri, Emanuela Salvi, Stefania Spila Alegiani (National Centre for Drug Research and Evaluation, National Institute of Health - Istituto Superiore di Sanità); Anna Rosa Marra, Patrizia Felicetti, Pasquale Marchione, Fiorella Petronzelli, Giuseppe Pimpinella, Loriana Tartaglia (Department of post-marketing surveillance, Italian Medicine Agency - Agenzia Italiana del Farmaco); Patrizio Pezzotti, Antonino Bella, Massimo Fabiani, Matteo Spuri, Alberto Mateo Urdiales (Infectious Disease Department, National Institute of Health - Istituto Superiore di Sanità); Lorenza Ferrara, Luca Bolognesi, Lucia Favella (Piemonte Region); Giuseppe Monaco, Olivia Leoni, Michele Ercolanoni, Marco Lazzeretti (Lombardia Region); Gianluca Trifirò, Ugo Moretti, Giovanna Scroccaro, Paola Deambrosis, Giovanna Zanoni, Manuel Zorzi, Emanuela Bovo, Michele Tonon, Elena Vecchiato (Veneto Region); Paola Rossi, Cristina Zappetti, Sara Samez, Elena Clagnan (Friuli Venezia Giulia Region); Ester Sapigni, Aurora Puccini, Nazanin Morgheiseh (Emilia Romagna Region); Marco Tuccori, Rosa Gini, Giulia Hyeraci, Valentina Borsi (Toscana Region); Lorella Lombardozzi, Valeria Desiderio, Nadia Mores, Valeria Belleudi, Maria Balducci, Francesca Romana Poggi (Lazio Region); Annalisa Capuano, Ugo Trama, Massimo Di Gennaro, Roberta Giordana, Maria Grazia Fumo (Campania Region); Silvio Tafuri, Pasquale Stefanizzi, Domenica Ancona (Puglia Region).

## Footnotes

### Contributors

MM and SSA contributed equally as joint first authors. MM, SSA, CM, PM, PF, RDC, FP, ARM and FMI conceived the study question and designed the study. MM, SSA, MS, VB, FRP, ML, ME, EC and EB retrieved and prepared the data. MM, SSA, CM and MS carried out the analysis. MM, SSA, CM, MS, PM, PF, GT, RDC, FP, LT and FMI interpreted data. MM, SSA, CM, and FMI wrote the first draft of the manuscript. MM, SSA, CM, PM, PF, GT, RDC, FP, LT and FMI critically revised the paper. All authors contributed to subsequent drafts and interpretation of the findings and approved the final version of the manuscript. FMI, MM, SSA are manuscript’s guarantors. The corresponding author (CM) attests that all listed authors meet authorship criteria and that no others meeting the criteria have been omitted.

### Competing interests

All authors have no conflicts of interest that are directly relevant to the content of this article. All authors have completed the ICMJE uniform disclosure form at www.icmje.org/disclosure-of-interest/.

The lead authors (the manuscript’s guarantors) affirms that the manuscript is an honest, accurate, and transparent account of the study being reported; that no important aspects of the study have been omitted; and that any discrepancies from the study as planned have been explained.

### Dissemination to public communities

The study coordinators will continue to update the analyses based on these data sources. The study findings will also be disseminated by ISS, AIFA and Regional institutions web sites.

## References

1. Diaz GA, Parsons GT, Gering SK, Meier AR, Hutchinson IV, Ari Robicsek A. Myocarditis and pericarditis after vaccination for COVID-19. JAMA 2021;326(12):1210–1212. doi:10.1001/jama.2021.13443 pmid: 34347001

2. Marshall M, Ferguson ID, Lewis P, et al. Symptomatic acute myocarditis in 7 adolescents after Pfizer-BioNTech COVID-19 vaccination. Pediatrics 2021 Sep;148 (3):e2021052478. doi:10.1542/peds.2021-052478 pmid:34088762

3. Montgomery J, Ryan M, Engler R, et al. Myocarditis following immunization with mRNA COVID-19 vaccines in members of the US military. JAMA Cardiol 2021;6(10):1202–1206. doi:10.1001/jamacardio.2021.2833 pmid:34185045

4. Centers for Disease Control and Prevention. COVID-19 VaST Technical Report - May 17, 2021. 2021. Available from: https://www.cdc.gov/vaccines/acip/work-groups-vast/technical-report-2021-05-17.html

5. European Medicines Agency. Meeting highlights from the Pharmacovigilance Risk Assessment Committee (PRAC) 3-6 May 2021. 2021. https://www.ema.europa.eu/en/news/meeting-highlights-pharmacovigilance-risk-assessment-committee-prac-3-6-may-2021

6. SPEAC - Safety Platform for Emergency vACcines. SO2-D2.1.2 Priority List of COVID-19 Adverse events of special interest: Quarterly update December 2020. https://brightoncollaboration.us/wp-content/uploads/2021/01/SO2_D2.1.2_V1.2_COVID-19_AESI-update_V1.3.pdf

7. EMA/PRAC/234052/2021. Consideration on core requirements for RMPs of COVID19 vaccines. coreRMP19 guidance v2.0. 10 June 2021. https://www.ema.europa.eu/en/documents/other/consideration-core-requirements-rmps-covid-19-vaccines_en.pdf

8. COVID-19 subcommittee of the WHO Global Advisory Committee on Vaccine Safety (GACVS): updated guidance regarding myocarditis and pericarditis reported with COVID-19 mRNA vaccines 9 July 2021 Statement. 2021. Available from: https://www.who.int/news/item/09-07-2021-gacvs-guidance-myocarditis-pericarditis-covid-19-mrna-vaccines

9. EMA Safety Communication. Comirnaty and Spikevax: possible link to very rare cases of myocarditis and pericarditis. https://www.ema.europa.eu/en/news/comirnaty-spikevax-possible-link-very-rare-cases-myocarditis-pericarditis.

10. Paterlini M. Covid-19: Sweden, Norway, and Finland suspend use of Moderna vaccine in young people “as a precaution” BMJ 2021; 375 doi: https://doi.org/10.1136/bmj.n2477

11. Epi-phare. Myocardite et péricardite après la vaccination Covid-19. https://www.epi-phare.fr/rapports-detudes-et-publications/myocardite-pericardite-vaccination-covid19/

12. EMA/PRAC/683817/2021. PRAC recommendations on signals. Adopted at the 29 November-2 December 2021 PRAC meeting. https://www.ema.europa.eu/en/documents/prac-recommendation/prac-recommendations-signals-adopted-29-november-2-december-2021-prac-meeting_en.pdf

13. Barda N, Dagan N, Ben-Shlomo Y, et al. Safety of the BNT162b2 mRNA Covid-34432976 19 vaccine in a nationwide setting. N Engl J Med 2021 Sep;385(12):1078–1090. doi: 10.1056/NEJMoa2110475 pmid: 34432976

14. Dagan N, Barda N, Balicer RD. Adverse effects after BNT162b2 vaccine and SARS-CoV-2 infection, according to age and sex. N Engl J Med 2021 Dec;385(24):2299. doi:10.1056/NEJMc2115045 pmid: 34706169

15. Klein NP, Lewis N, Goddard K, et al. Surveillance for adverse events after COVID-19 mRNA vaccination. JAMA 2021;326(14):1390–1399. doi:10.1001/jama.2021.15072 pmid:34477808

16. Patone M, Mei XW, Handunnetthi L, et al. Risks of myocarditis, pericarditis, and cardiac arrhythmias associated with COVID-19 vaccination or SARS-CoV-2 infection. Nat Med Dec;2021. doi:10.1038/s41591-021-01630-0 pmid:34907393

17. Patone M, Mei X W, Handunnetthi L, et al. Risk of myocarditis following sequential COVID-19 vaccinations by age and sex. MedRxiv 2021.12.23.21268276 [Preprint]. 2021 [cited 2022 January 21]. Available from: https://www.medrxiv.org/content/10.1101/2021.12.23.21268276v1

18. Husby A, Hansen JV, Fosbøl E, et al. SARS-CoV-2 vaccination and myocarditis or myopericarditis: population based cohort study. BMJ 2021;375:e068665. doi:10.1136/bmj-2021-068665 pmid:34916207

19. Massari M, Spila Alegiani S, Da Cas R, Menniti Ippolito F. TheShinISS: an open-source tool for conducting distributed analyses within pharmacoepidemiological multi-database studies. Boll Epidemiol Naz 2020;1(2):39–45. doi:10.53225/BEN_006

20. Whitaker HJ, Farrington CP, Spiessens B, Musonda P. Tutorial in bio-statistics: the self-controlled case series method. Stat Med 2006;25(10):1768–97. doi:10.1002/sim.2302 pmid: 16220518

21. Petersen I, Douglas I, Whitaker H. Self controlled case series methods: an alternative to standard epidemiological study designs. BMJ 2016 Sep;354:i4515. doi:10.1136/bmj.i4515. pmid:27618829

22. Weldeselassie YG, Whitaker HJ, Farrington CP. Use of the self-controlled case-series method in vaccine safety studies: review and recommendations for best practice. Epidemiol Infect 2011;139 (12):1805–17. doi:10.1017/S0950268811001531 pmid:21849099

23. Farrington CP, Whitaker HJ, Hocine MN. Case series analysis for censored, perturbed, or curtailed post-event exposures. Biostatistics 2009;10(1):3–16. doi:10.1093/biostatistics/kxn013 pmid:18499654

24. Farrington CP, Whitaker H, Weldeselassie YG. Self-Controlled Case Series Studies. A Modelling Guide with R. CRC Press, 2018

25. Galeotti F, Massari M, D’Alessandro R, et al. Risk of Guillain-Barré syndrome after 2010-2011 influenza vaccination. Eur J Epidemiol 2013 May;28(5):433–44. doi: 10.1007/s10654-013-9797-8. Epub 2013 Mar 31. pmid:23543123

26. Trifirò G, Massari M, Da Cas R, et al. RAAS inhibitor group. Renin–Angiotensin–Aldosterone System Inhibitors and Risk of Death in Patients Hospitalised with COVID19: A Retrospective Italian Cohort Study of 43,000 Patients. Drug Saf 2020 Dec; 43(12):1297–1308. doi:10.1007/s40264-020-00994-5 pmid: 32852721

27. Trifirò G, Isgrò V, Ingrasciotta Y, et al. Large-Scale Postmarketing Surveillance of Biological Drugs for Immune-Mediated Inflammatory Diseases Through an Italian Distributed Multi-Database Healthcare Network: The VALORE Project. BioDrugs 2021 Nov;35(6):749–764. doi:10.1007/s40259-021-00498-3. pmid:34637126

28. Rosa AC, Marino ML, Finocchietti M, et al. Immunosuppressive therapy after solid organ transplantation in Italy: a pilot study of the CESIT* project. XI Congresso Nazionale SISMEC; 2021 Sep 15-18; Bari,IT. https://sismecbari2021.it/wp-content/uploads/2021/09/39-ROSA-ALESSANDRO.pdf

29. Wilson K. and Hawken S. Drug safety studies and measures of effect using the self-controlled case series design. Pharmacoepidemiol Drug Saf 2013;22:108–110. doi:10.1002/pds.3337 pmid:22915354

30. COVID-19 subcommittee of the WHO Global Advisory Committee on Vaccine Safety (GACVS) reviews cases of mild myocarditis reported with COVID-19 mRNA vaccines GACVS 26 May 2021. 2021. Available from: https://www.who.int/news/item/26-05-2021-gacvs-myocarditis-reported-with-covid-19-mrna-vaccines

31. Weldeselassie YJ, Whitaker H and Farrington P (2021). SCCS: The Self-Controlled Case Series Method. R. package version 1.5. Available from: https://CRAN.R-project.org/package=SCCS

32. Mei R, Raschi E, Forcesi E, et al. Myocarditis and pericarditis after immunization: gaining insights through the vaccine adverse event reporting system. Int J Cardiol. 2018; 273: 183–186.

33. Vojdani A, Kharrazian D. Potential antigenic cross-reactivity between SARS-CoV-2 and human tissue with a possible link to an increase in autoimmune diseases. Clin Immunol. 2020 Aug; 217():108480.

34. Caso F, Costa L, Ruscitti P, et al. Could Sars-coronavirus-2 trigger autoimmune and/or autoinflammatory mechanisms in genetically predisposed subjects? Autoimmun Rev. 2020 May; 19(5):102524

35. Fox SE, Falgout L, Vander Heide RS. COVID-19 Myocarditis: Quantitative analysis of the inflammatory infiltrate and a proposed mechanism. Cardiovascular Pathology. Cardiovascular Pathology. 2021; 54: 107361

36. Engler RJM, Nelson MR, LC Collins Jr., et al. A Prospective Study of the Incidence of myocarditis/Pericarditis and New Onset Cardiac Symptoms following Smallpox and Influenza Vaccination. PLOS ONE. 2015;10(3):e0118283. https://doi.org/10.1371/journal.pone.0118283

37. Grimaud M, Starck J, Levy M, et al. Acute myocarditis and multisystem inflammatory emerging disease following SARS-CoV-2 infection in critically ill children. Ann Intensive Care. 2020 Jun 1; 10(1):69.

38. Fairweather D, Cooper LT, Blauwet LA. Sex and Gender Differences in Myocarditis and Dilated Cardiomyopathy. Curr Probl Cardiol. 2013 January; 38(1): 7–46

39. Clerkin KJ, Fried JA, Raikhelkar J, et al. COVID-19 and cardiovascular disease. Circulation. 2020;141:1648–1655.

40. Bozkurt B, Kamat I, Hotez P. Myocarditis With COVID-19 mRNA Vaccines. Circulation 2021;144:471–484.

