## Supplemental material for "Post-marketing active surveillance of myocarditis and pericarditis following vaccination with COVID-19 mRNA vaccines in persons aged 12-39 years in Italy: a multi-database, self-controlled case series study"

**Figure S1**- Diagram showing the data flow when using *TheShinISS* to locally elaborate health care data structured according to a Common Data Model.

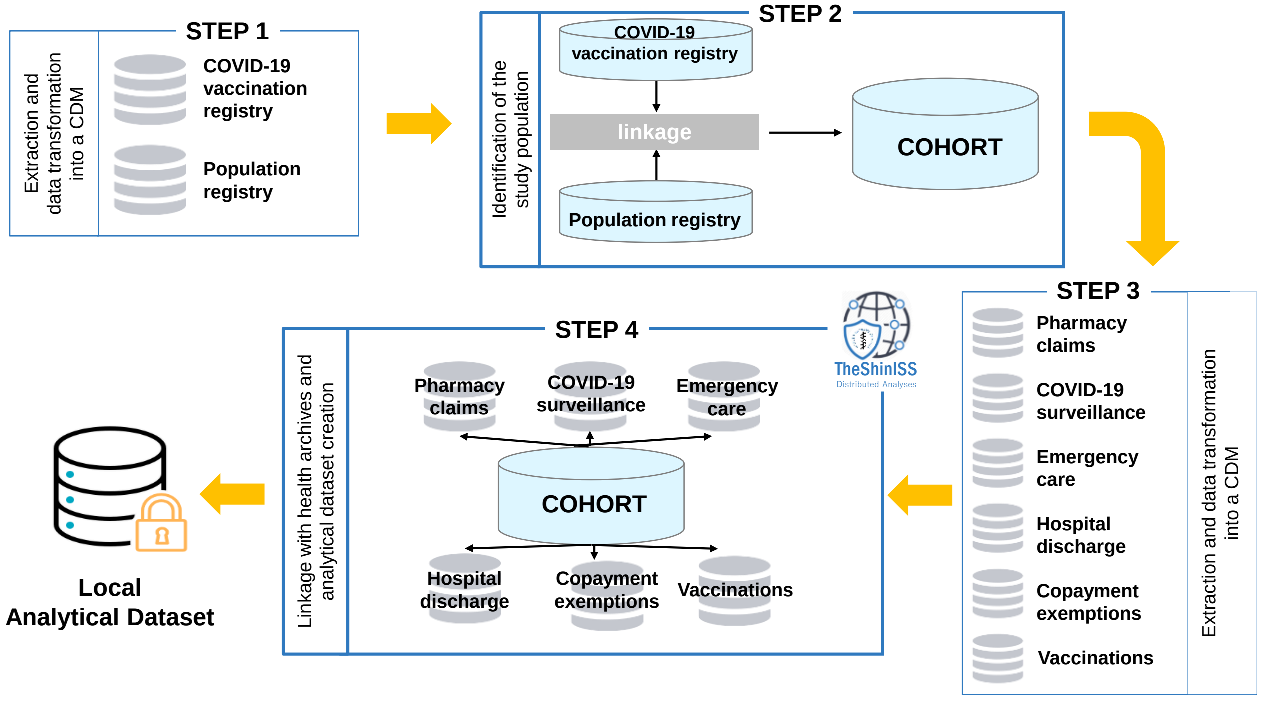


CDM: Common Data Model

**Table S1.** Definition of study comorbidities.

| Comorbidities | Databases | | |
|---|---|---|---|
| | Hospital discharge | Pharmacy claims | Copayment exemption |
| | ICD 9-CM codes (in the last 5 years) | ATC codes (in the last 12 months) | Exemption codes |
| COPD | 490; 492; 493; 494; 496 | R03 | 057; 007 |
| Chronic pulmonary disease | 480-488; 491; 495; 518.81-518.84 | J05AH | 024 |
| Chronic kidney failure | 580; 582-585; 593; 753.12-753.14 | | 023; 022; 061; 062 |
| Neoplasms | 140-209, V10 | L01 | 048 |
| Diabetes mellitus | 250 | A10 | 013 |
| Hematologic disease | 280-284; 285 (excl.285.1); 286-289 | B01AA; B01AB; B01AE; B01AF; B01AX; B02BD; B03 | 003 |
| Cardiovascular and cerebrovascular diseases | 390-398; 406-459 | B01AC; C01B; C01DA; C08DA; C08DB | 002; 021 0A02; 0B02; 0C02; 036 |
| Hypertension | 401-405 | C02; C03; C07; C08 (excl. C08DA; C08DB); C09 | 031; 0A31; 0031 |
| Hepatopathy | 456.0-456.2; 571-572; 573.0 | | 008; 016 |
| Dementia /Alzheimer | 290; 294.1; 331.2 | N06DA, N06DX | 011; 029 |
| HIV | 042 | J05AE; J05AF; J05AG; J05AR | 020 |
| Rheumatic diseases | 446.5; 710; 714; 720; 725; 696 | L04 | 006;028; 030; 045; 054; 067 |
| Cystic fibrosis | 277.0 | R07AX | 018 |
| Neurological diseases | 296.3; 238.7; 311; 332; 340; 345; 348.39 | N03A; N04B; N05A; N06A | 017; 038; 044; 046 |
| Peptic Ulcer | 531-533 | A02B | |
| Colitis | 555; 556 | | 009 |
| Celiac disease | 579.0 | | 059 |
| Infection (in the last 12 months) | 053; 599.0; 010-018; 031; 078.5; 052-054; 136.3; 117.5 | J01; J02; J04; J05 (excl. J05AE; J05AF; J05AG; J05AR; J05AH) | 055 |
| Corticosteroids for systemic use | | H02A | |
| NSAIDs use | | M01A | |
| Estroprogestinics use | | G03 | |

ICD: International Classification of Disease; ATC: Anatomical Therapeutic Chemical Classification System; COPD: Chronic obstructive pulmonary disease; HIV: Human Immunodeficiency Virus; NSAIDs: Non-steroidal anti-inflammatory drugs

**Table S2** - Relative incidence estimated by adjusted SCCS and excess cases per 100,000 vaccinated by risk intervals: 232 myocarditis and/or pericarditis events in the BNT162b2 vaccinated males aged 12-39 years from 27 December 2020 to 30 September 2021.

| **Risk interval** | **Dose** | **Events in the risk interval (n)** | **Adjusted Relative Incidence (95% CI)*** | **Excess cases per 100,000 vaccinated (95% CI)**** |
|---|---|---|---|---|
| [0-7) | Dose 1 | 9 | 1.53 (0.71-3.31) | |
| | Dose 2 | 13 | 3.45 (1.78-6.68) | 1.0 (0.6-1.2) |
| [7-14) | Dose 1 | 9 | 1.53 (0.74-3.17) | |
| | Dose 2 | 4 | 1.01 (0.37-2.75) | |
| [14-21) | Dose 1 | 8 | 1.41 (0.63-3.14) | |
| | Dose 2 | 5 | 1.29 (0.49-3.42) | |
| *Ref.* | | *184* | *1.0* | |

SCCS: Self-Controlled Cases Series; n.: number; CI: Confidence interval; Ref.: reference period (unexposed period). *adjusted by calendar period; **excess cases are not given when RI did not show a statistically significant increase of incidence over the [0-21) risk interval

**Table S3** - Relative incidence estimated by adjusted SCCS and excess cases per 100,000 vaccinated by risk intervals: 114 myocarditis and/or pericarditis events in the BNT162b2 vaccinated females aged 12-39 years from 27 December 2020 to 30 September 2021.

| **Risk interval** | **Dose** | **Events in the risk interval (n)** | **Adjusted Relative Incidence (95% CI)*** | **Excess cases per 100,000 vaccinated (95% CI)**** |
|---|---|---|---|---|
| [0-7) | Dose 1 | 5 | 0.88 (0.34-2.32) | |
| | Dose 2 | 9 | 3.38 (1.47-7.74) | 0.7 (0.3-0.9) |
| [7-14) | Dose 1 | 1 | 0.19 (0.02-1.63) | |
| | Dose 2 | 3 | 1.16 (0.35-3.85) | |
| [14-21) | Dose 1 | 3 | 0.71 (0.21-2.44) | |
| | Dose 2 | 5 | 2.01 (0.72-5.62) | |
| *Ref.* | | *88* | *1.0* | |

SCCS: Self-Controlled Cases Series; n.: number; CI: Confidence interval; Ref.: reference period (unexposed period). *adjusted by calendar period; **excess cases are not given when RI did not show a statistically significant increase of incidence over the [0-21) risk interval

**Table S4** - Relative incidence estimated by adjusted SCCS and excess cases per 100,000 vaccinated by risk intervals: 46 myocarditis and/or pericarditis events in the BNT162b2 vaccinated population aged 12-17 years from 27 December 2020 to 30 September 2021.

| Risk interval | Dose | Events in the risk interval (n) | Adjusted Relative Incidence (95% CI)* | Excess cases per 100,000 vaccinated (95% CI)** |
|---|---|---|---|---|
| [0-7) | Dose 1 | 3 | 1.06 (0.17-6.59) | |
| | Dose 2 | 7 | 5.74 (1.52-21.72) | 1.7 (0.7-1.9) |
| [7-14) | Dose 1 | 3 | 0.99 (0.24-4.19) | |
| | Dose 2 | 1 | 0.74 (0.11-5.15) | |
| [14-21) | Dose 1 | 1 | 0.36 (0.05-2.72) | |
| | Dose 2 | 0 | — | |
| *Ref.* | | *31* | *1.0* | |

SCCS: Self-Controlled Cases Series; n.: number; CI: Confidence interval; Ref.: reference period (unexposed period). *adjusted by calendar period; **excess cases are not given when RI did not show a statistically significant increase of incidence over the [0-21) risk interval

**Table S5** - Relative incidence estimated by adjusted SCCS and excess cases per 100,000 vaccinated by risk intervals: 154 myocarditis and/or pericarditis events in the BNT162b2 vaccinated population aged 18-29 years from 27 December 2020 to 30 September 2021.

| Risk interval | Dose | Events in the risk interval (n) | Adjusted Relative Incidence (95% CI)* | Excess cases per 100,000 vaccinated (95% CI)** |
|---|---|---|---|---|
| [0-7) | Dose 1 | 7 | 1.76 (0.76-4.05) | |
| | Dose 2 | 11 | 4.02 (1.81-8.91) | 1.1 (0.6-1.3) |
| [7-14) | Dose 1 | 3 | 0.72 (0.20-2.61) | |
| | Dose 2 | 4 | 1.46 (0.51-4.14) | |
| [14-21) | Dose 1 | 4 | 1.00 (0.34-2.96) | |
| | Dose 2 | 4 | 1.56 (0.55-4.44) | |
| *Ref.* | | *121* | *1.0* | |

SCCS: Self-Controlled Cases Series; n.: number; CI: Confidence interval; Ref.: reference period (unexposed period). *adjusted by calendar period; **excess cases are not given when RI did not show a statistically significant increase of incidence over the [0-21) risk interval

**Table S6**- Relative incidence estimated by adjusted SCCS and excess cases per 100,000 vaccinated by risk intervals: 146 myocarditis and/or pericarditis events in the BNT162b2 vaccinated population aged 30-39 years from 27 December 2020 to 30 September 2021.

| Risk interval | Dose | Events in the risk interval (n) | Adjusted Relative Incidence (95% CI)* | Excess cases per 100,000 vaccinated (95% CI)** |
|---|---|---|---|---|
| [0-7) | Dose 1 | 4 | 0.86 (0.31-2.38) | |
| | Dose 2 | 4 | 1.64 (0.59-4.53) | |
| [7-14) | Dose 1 | 4 | 0.97 (0.34-2.80) | |
| | Dose 2 | 2 | 0.84 (0.21-3.38) | |
| [14-21) | Dose 1 | 6 | 1.68 (0.70-4.04) | |
| | Dose 2 | 6 | 2.50 (0.97-6.45) | |
| *Ref.* | | *120* | *1.0* | |

SCCS: Self-Controlled Cases Series; n.: number; CI: Confidence interval; Ref.: reference period (unexposed period). *adjusted by calendar period; **excess cases are not given when RI did not show a statistically significant increase of incidence over the [0-21) risk interval

**Table S7** - Relative incidence estimated by adjusted SCCS and excess cases per 100,000 vaccinated by risk intervals: 70 myocarditis and/or pericarditis events in the mRNA-1273 vaccinated males aged 12-39 years from 27 December 2020 to 30 September 2021.

| Risk interval | Dose | Events in the risk interval (n) | Adjusted Relative Incidence (95% CI)* | Excess cases per 100,000 vaccinated (95% CI)** |
|---|---|---|---|---|
| [0-7) | Dose 1 | 10 | 12.28 (4.09-36.83) | 3.8 (3.1-4.0) |
| | Dose 2 | 19 | 11.91 (3.88-36.53) | 8.8 (7.2-9.4) |
| [7-14) | Dose 1 | 2 | 2.19 (0.42-11.41) | |
| | Dose 2 | 0 | — | |
| [14-21) | Dose 1 | 0 | — | |
| | Dose 2 | 1 | 0.65 (0.08-5.37) | |
| *Ref.* | | *38* | *1.0* | |

SCCS: Self-Controlled Cases Series; n.: number; CI: Confidence interval; Ref.: reference period (unexposed period). *adjusted by calendar period; **excess cases are not given when RI did not show a statistically significant increase of incidence over the [0-21) risk interval

**Table S8** - Relative incidence estimated by adjusted SCCS and excess cases per 100,000 vaccinated by risk intervals: 25 myocarditis and/or pericarditis events in the mRNA-1273 vaccinated females aged 12-39 years from 27 December 2020 to 30 September 2021.

| Risk interval | Dose | Events in the risk interval (n) | Adjusted Relative Incidence (95% CI)* | Excess cases per 100,000 vaccinated (95% CI)** |
|---|---|---|---|---|
| [0-7) | Dose 1 | 1 | 0.69 (0.08-5.75) | |
| | Dose 2 | 4 | 2.08 (0.45-9.72) | |
| [7-14) | Dose 1 | 1 | 0.69 (0.10-4.87) | |
| | Dose 2 | 0 | — | |
| [14-21) | Dose 1 | 1 | 0.69 (0.07-6.86) | |
| | Dose 2 | 1 | 0.72 (0.13-4.05) | |
| *Ref.* | | *17* | *1.0* | |

SCCS: Self-Controlled Cases Series; n.: number; CI: Confidence interval; Ref.: reference period (unexposed period). *adjusted by calendar period; **excess cases are not given when RI did not show a statistically significant increase of incidence over the [0-21) risk interval

**Table S9** - Relative incidence estimated by adjusted SCCS and excess cases per 100,000 vaccinated by risk intervals: 11 myocarditis and/or pericarditis events in the mRNA-1273 vaccinated population aged 12-17 years from 27 December 2020 to 30 September 2021.

| Risk interval | Dose | Events in the risk interval (n) | Adjusted Relative Incidence (95% CI)*§ | Excess cases per 100,000 vaccinated (95% CI)** |
|---|---|---|---|---|
| [0-7) | Dose 1 | 0 | | |
| | Dose 2 | 3 | | |
| [7-14) | Dose 1 | 0 | | |
| | Dose 2 | 0 | | |
| [14-21) | Dose 1 | 0 | | |
| | Dose 2 | 1 | | |
| *Ref.* | | *7* | | |

SCCS: Self-Controlled Cases Series; n.: number; CI: Confidence interval; Ref.: reference period (unexposed period). *adjusted by calendar period; **excess cases are not given when RI did not show a statistically significant increase of incidence over the [0-21) risk interval; §considering the small number of cases in this age group, it was not possible to provide any estimates.

**Table S10** - Relative incidence estimated by adjusted SCCS and excess cases per 100,000 vaccinated by risk intervals: 57 myocarditis and/or pericarditis events in the mRNA-1273 vaccinated population aged 18-29 years from 27 December 2020 to 30 September 2021.

| Risk interval | Dose | Events in the risk interval (n) | Adjusted Relative Incidence (95% CI)* | Excess cases per 100,000 vaccinated (95% CI)** |
|---|---|---|---|---|
| [0-7) | Dose 1 | 9 | 7.58 (2.62-21.94) | 3.5 (2.4-3.8) |
| | Dose 2 | 18 | 9.58 (3.32-27.58) | 8.3 (6.7-9.2) |
| [7-14) | Dose 1 | 1 | 0.78 (0.09-6.51) | |
| | Dose 2 | 0 | — | |
| [14-21) | Dose 1 | 0 | — | |
| | Dose 2 | 1 | 0.55 (0.07-4.37) | |
| *Ref.* | | *28* | *1.0* | |

SCCS: Self-Controlled Cases Series; n.: number; CI: Confidence interval; Ref.: reference period (unexposed period). *adjusted by calendar period; **excess cases are not given when RI did not show a statistically significant increase of incidence over the [0-21) risk interval

**Table S11** - Relative incidence estimated by adjusted SCCS and excess cases per 100,000 vaccinated by risk intervals: 27 myocarditis and/or pericarditis events in the mRNA-1273 vaccinated population aged 30-39 years from 27 December 2020 to 30 September 2021.

| Risk interval | Dose | Events in the risk interval (n) | Adjusted Relative Incidence (95% CI)* | Excess cases per 100,000 vaccinated (95% CI)** |
|---|---|---|---|---|
| [0-7) | Dose 1 | 2 | 6.57 (1.32-32.63) | 1.0 (0.3-1.1) |
| | Dose 2 | 2 | 3.22 (0.69-15.10) | |
| [7-14) | Dose 1 | 2 | 5.87 (1.34-25.74) | 1.0 (0.3-1.1) |
| | Dose 2 | 0 | — | |
| [14-21) | Dose 1 | 1 | 2.56 (0.23-28.57) | |
| | Dose 2 | 0 | — | |
| *Ref.* | | *20* | *1.0* | |

SCCS: Self-Controlled Cases Series; n.: number; CI: Confidence interval; Ref.: reference period (unexposed period). *adjusted by calendar period; **excess cases are not given when RI did not show a statistically significant increase of incidence over the [0-21) risk interval

**Figure S2**- Relative Incidence in the [0-7) risk period after mRNA vaccination in the vaccinated population aged 12-39 years from 27 December 2020 to 30 September 2021 by vaccine brand, gender and age group.

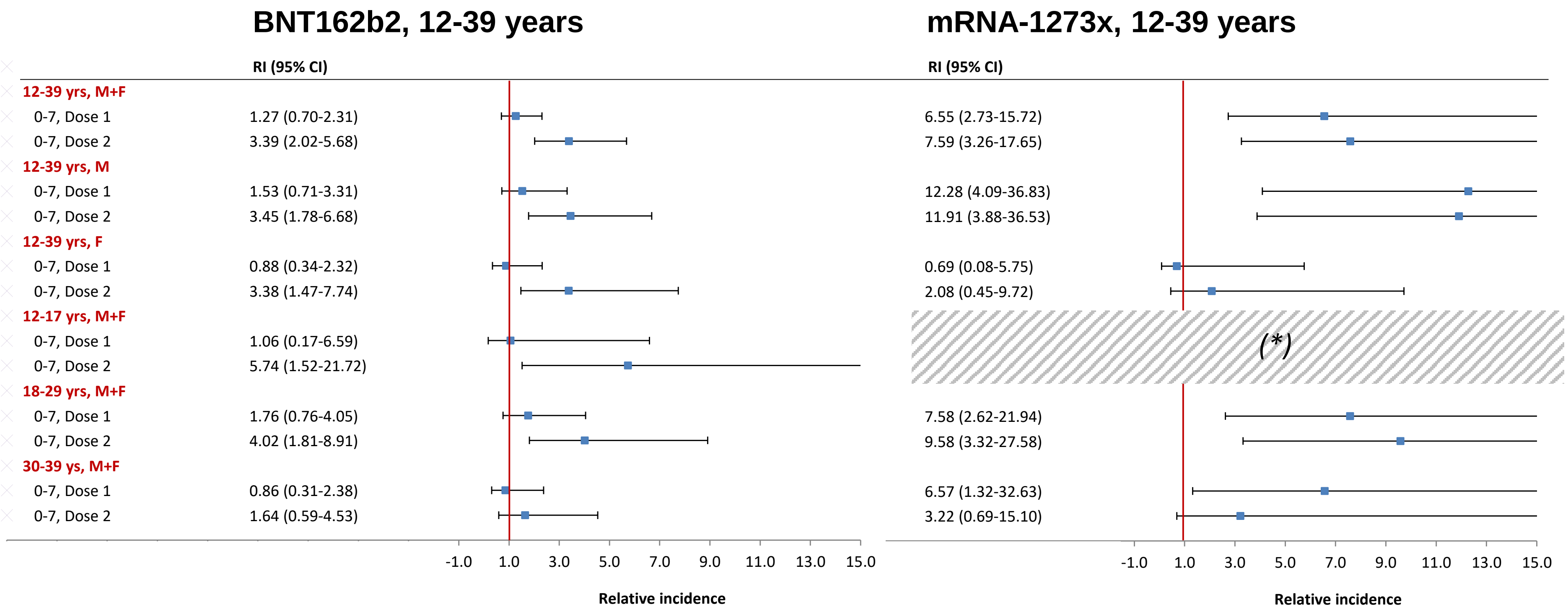


RI: Relative Incidence; CI: Confidence Interval; M: Males; F: Females. (*) Considering the small number of cases in the vaccinated with mRNA-1273 of age 12-17 years, it was not possible to provide any estimates

**Table S12** – Sensitivity analyses.

| | Risk interval | Dose | Adjusted Relative Incidence (95% CI)* | |
|---|---|---|---|---|
| | | | BNT162b2 | mRNA-1273 |
| **Primary analysis** | **[0-7)** | **Dose 1** | **1.27 (0.70-2.31)** | **6.55 (2.73-15.72)** |
| | | **Dose 2** | **3.39 (2.02-5.68)** | **7.59 (3.26-17.65)** |
| **a) SCCS standard** SCSS standard cutting the observation time at the first and second dose (n. 441) | **[0-7)** | Dose 1 | 1.27 (0.69-2.33) | 5.59 (2.37-13.17) |
| | | Dose 2 | 3.35 (1.98-5.69) | 6.20 (2.45-15.68) |
| **b) Observation time period (I)** analysis restricted to the study period from 27 December 2020 to 31 May 2021 (n. 85)** | **[0-7)** | Dose 1 | 0.48 (0.10-2.20) | *** |
| | | Dose 2 | 3.29 (1.22-8.88) | *** |
| **b) Observation time period (II)** analysis restricted to the study period from 1 June 2021 to 30 September 2021 (n. 160)** | **[0-7)** | Dose 1 | 2.05 (0.97-4.33) | 6.26 (2.15-15.22) |
| | | Dose 2 | 4.32 (2.11-8.83) | 13.43 (4.65-38.76) |
| **b) Exposure time period** excluding day 0 from [0-7) day risk interval (n. 441) | **[0-7)** | Dose 1 | 1.39 (0.75-2.55) | 7.68 (3.21-18.39) |
| | | Dose 2 | 3.96 (2.36-6.62) | 8.76 (3.78-20.29) |
| **c) Heterologous vaccination (I)** primary analysis excluding individuals with two different brands in the first and second dose (n. 440) | **[0-7)** | Dose 1 | 1.27 (0.69-2.31) | 6.56 (2.73-15.74) |
| | | Dose 2 | 3.39 (2.02-5.68) | 7.59 (3.26-17.66) |
| **c) Heterologous vaccination (II)** censoring 1 subjects who received a different brand on the II dose (from mRNA-1273 to BNT162b2) | **[0-7)** | Dose 1 | *** | 6.61 (2.75-18.90) |
| | | Dose 2 | | 7.61 (3.27-17.72) |
| **d) SARS-CoV-2 infection** restricting the analyses to subjects without a SARS-CoV-2 positive test before and during the study period (n. 378) | **[0-7)** | Dose 1 | 1.17 (0.64-2.25) | 5.66 (1.99-16.13) |
| | | Dose 2 | 3.13 (1.82-5.39) | 7.85 (3.26.18.94) |

SCCS: Self-Controlled Cases Series; CI: Confidence interval. *adjusted by calendar period; **subjects vaccinated and with event in each period were included in this analysis ***considering the small number of cases, it was not possible to provide any estimates

**Table S13** – - Relative incidence estimated by adjusted SCCS and excess cases per 100,000 vaccinated by risk intervals: 1,759 myocarditis and/or pericarditis events in the BNT162b2 and 291 events in the mRNA-1273 vaccinated population aged ≥40 years from 27 December 2020 to 30 September 2021 (ancillary analysis).

| **Gender** | **Risk interval** | **Dose** | **BNT162b2 Vaccinated 6,835,634 (n. cases 1,759)** | | | **mRNA-1273 Vaccinated 1,041,582 (n. cases 291)** | | |
|---|---|---|---|---|---|---|---|---|
| | | | **Events in the risk interval (n)** | **Adjusted Relative Incidence (95% CI)*** | **Excess cases per 100,000 Vaccinated (95% CI)**** | **Events in the risk interval (n)** | **Adjusted Relative Incidence (95% CI)*** | **Excess cases per 100,000 Vaccinated (95% CI)**** |
| **Males+Females** | **[0-7)** | Dose 1 | 39 | 0.59 (0.42-0.82) | | 7 | 0.56 (0.23-1.36) | |
| | | Dose 2 | 43 | 0.84 (0.61-1.16) | | 10 | 1.11 (0.57-2.17) | |
| | | *Ref.* | *1,479* | *1* | | *247* | *1* | |
| **Males** | **[0-7)** | Dose 1 | 16 | 0.43 (0.26-0.71) | | 1 | 0.12 (0.01-1.31) | |
| | | Dose 2 | 23 | 0.85 (0.54-1.33) | | 6 | 1.11 (0.47-2.66) | |
| | | *Ref.* | *824* | *1* | | *134* | *1* | |
| **Females** | **[0-7)** | Dose 1 | 23 | 0.76 (0.49-1.19) | | 6 | 1.16 (0.46-2.92) | |
| | | Dose 2 | 20 | 0.84 (0.53-1.33) | | 4 | 1.11 (0.39-3.13) | |
| | | *Ref.* | *655* | *1* | | *113* | *1* | |

SCCS: Self-Controlled Cases Series; n.: number; CI: Confidence interval; Ref.: reference period (unexposed period). *adjusted by calendar period; **excess cases are not given when RI did not show a significant increase of incidence over the [0-21) risk interval
